# COVID-19 susceptibility variants associate with blood clots, thrombophlebitis and circulatory diseases

**DOI:** 10.1101/2021.05.04.21256617

**Authors:** Areti Papadopoulou, Hanan Musa, Mathura Sivaganesan, David McCoy, Panos Deloukas, Eirini Marouli

## Abstract

**Objective:** Epidemiological studies suggest that individuals with comorbid conditions including diabetes, chronic lung diseases and others, are at higher risk of adverse COVID-19 outcomes. Genome-wide association studies have identified several loci associated with increased susceptibility and severity for COVID-19. However, it is not clear whether these associations are genetically determined or not. We used a Phenome-Wide Association (PheWAS) approach to investigate the role of genetically determined COVID-19 susceptibility on disease related outcomes.

**Approach and Results:** PheWAS analyses were performed in order to identify traits and diseases related to COVID-19 susceptibility and severity, evaluated through a predictive COVID-19 risk score. We utilised phenotypic data in up to 400,000 individuals from the UK Biobank, including Hospital Episode Statistics and General Practice data. We identified a spectrum of associations between both genetically determined COVID-19 susceptibility and severity with a number of traits. COVID-19 risk was associated with increased risk for phlebitis and thrombophlebitis (OR = 1.11, p = 5.36e^-08^). We also identified significant signals between COVID-19 susceptibility with blood clots in the leg (OR= 1.1, p = 1.66e^-16^) and with increased risk for blood clots in the lung (OR = 1.12, p = 1.45 e^-10^).

**Conclusions:** Our study identifies significant association of genetically determined COVID-19 with increased blood clot events in leg and lungs. The reported associations between both COVID-19 susceptibility and severity and other diseases adds to the identification and stratification of individuals at increased risk, adverse outcomes and long-term effects.

## Introduction

Corona Virus Disease 2019 (COVID-19) pertains to the disease caused by the virus SARS-CoV-2 - severe acute respiratory syndrome coronavirus 2. This RNA virus belongs to a family of coronaviruses that includes Middle East Respiratory Syndrome (MERS) and Severe Acute Respiratory Syndrome (SARS) [1]. COVID-19 was declared a pandemic by the WHO in March 2020, and as of the 16th of April 2021, there have been 139,802,523 confirmed cases documented in 219 countries [2].

Upon SARS-CoV-2 infection, typical symptoms include dry cough, fever, and loss of smell and/or taste. However, the presentation of infection can range from asymptomatic to critical illness which has led to categorisation of disease based on severity of symptoms. One definition of severe COVID-19 is the need for admission to an intensive care unit and need for a ventilator/supplementary oxygen which is less likely in mild to moderate COVID-19. The need for hospitalisation is mainly due to infection of the lower respiratory tract and therefore inflammation of lung alveoli, also known as pneumonia. This can lead to acute respiratory distress syndrome (ARDS), sepsis and multi-organ failure which are some of the main causes of mortality in COVID patients [3]. Aside from classifying severity of disease based on symptoms, histopathological differences can also be used for classification purposes. For example, it has been suggested that the formation of hyaline membranes, due to lack of surfactant leading to alveolar collapse, are characteristic of pneumonia and therefore of severe COVID-19 [3].

The development of severe COVID is attributed to a dysregulated immune response characterised by a cytokine storm, a severe and sustained activation of the inflammatory pathway mediated by overexpression of cytokines[4]. Cytokines usually act as a signal for innate immune cells, such as neutrophils and macrophages, to move to the site of infection. However, when this is inappropriately sustained, the increased vascular permeability can lead to pulmonary consolidation and other lung tissue injury. In addition to accumulation of pulmonary exudates, the signalling cascade can result in apoptosis of lung epithelial cells which can lead to the complication of ARDS since oxygen transfer is severely impaired. Studies have reported a correlation of higher levels of pro-inflammatory cytokines (e.g. IL-6) and inflammatory markers (e.g. C-reactive protein (CRP)) with more severe clinical presentations [3]. There is also evidence that type 1 interferon, an antiviral defence, is suppressed during the initial phase of the disease, which promotes the over-activity of the inflammatory pathway underlying disease severity [5]. The combination of low interferon levels and high pro-inflammatory cytokines is what indicates the dysregulated innate immune response to COVID.

Following genome-wide association studies (GWAS)of susceptibility to severe COVID infection, Mendelian Randomisation (MR) can then be used for causal inference of the role of a specific variant or risk factor. The outputs of such analyses could serve as guide in clinical trials aiming to repurpose existing medications for effective COVID management. Baricitinib is an example of an anti-inflammatory rheumatoid arthritis drug that has been repurposed to help buffer the cytokine storm that leads to lung injury [6]. Furthermore, evidence of under expression of IFNAR2, which encodes the receptor of interferon, amongst severe COVID patients has led to clinical trials of interferons to reduce mortality rate [4]. In addition, genomic analyses could reveal new therapeutic targets for drug development. 3CLpro, PLpro and RdRp, the main enzymes responsible for SARS-COV-2 replication using host machinery, emerged as potential therapeutic targets [7].

GWAS have identified several loci associated with increased susceptibility to COVID-19 infection and severe disease [8]. The underlying genes of these association signals can be broadly divided into underactive genes that enable viral replication and overactive genes that result in pulmonary inflammation and the symptoms of severe COVID-19. The most prominent findings include reduced activity of IFNAR2, increased activity of TYK2 and mutation of OAS1 which means poor activation of an enzyme involved in stopping viral replication [9].

An additional finding is a gene cluster on chromosome 3 that is associated with severe COVID-19 outcomes. COVID-19 individuals with gene variants at this locus had 1.6 odds of requiring hospitalisation [10]. The same study found that the susceptibility haplotype was most strongly associated with people of Bangladeshi origin which may in part explain the higher COVID-19 related mortality amongst Bangladeshi men compared to White males [11]. It is theorised that historically, gene flow from Neanderthals in this part of the world may have provided protection against certain pathogens through selection pressure amongst people who live in or come from what is now known as Bangladesh, but in the case of COVID-19 it confers a greater risk of severe infection [10]. The COVID-19 susceptibility haplotype showed similarity to Neanderthal Vindija haplotypes [10].

The risk of infection and severity of COVID-19 is modulated by various risk factors. The most frequently reported finding in the literature is that older patients or those with existing respiratory or cardiovascular disease are more susceptible to severe infection. A study found that a 60–74-year-old individual has a 5-times higher risk of hospitalisation and 90 times higher risk of death from COVID-19 compared to an 18–29-year-old [12]. The other most significant risk factor is body mass index (BMI) [13], which has a biological basis since obesity is associated with other conditions such as heart disease and diabetes, both of which are independent risk factors. Genetic susceptibility to obesity was found in MR studies to be associated with increased risk of infection [13]. The authors suggest that obesity could be causally linked to infection because of impaired lung function, increased baseline adipokines and cytokines which increase risk to developing ARDs. Additional studies support the causal relationship between obesity as a risk factor, although some findings suggest that this relationship might be mediated by type 2 diabetes [14].

Cardiovascular disease is also a predictor of poor COVID-19 outcomes. The combination of the hyperinflammatory environment due to a SARS-CoV-2 infection also puts the patient into a hypercoagulable state which leads to complications such as multi-organ failure, observed complication of severe COVID-19 [15]. Furthermore, hypertension could be a causal risk factor due to the viral spike protein binding to ACE2 receptors, thereby enhancing the activity of the renin-angiotensin-aldosterone system which leads to inflammation and fibrosis [16]. Similarly, smoking is a risk factor that has been found to increase expression of the ACE2 receptor in the lung which provides some explanation for why smokers are more likely to require hospital admission [17].

In the present study we aim to identify traits and diseases that are related to the genetic susceptibility for COVID-19. We conducted Phenome-Wide Association Analyses to evaluate the association of genetically determined COVID-19 susceptibility and severity using Predictive Risk Scores (PRS) with a comprehensive set of phenotypes including disease in the UK Biobank. COVID-19-related outcomes were derived from the COVID-19 Host Genetics Initiative (https://www.covid19hg.org/).

## Methods

Phenome-wide association studies (PheWAS) are used to identify the effects of a trait of interest across a potentially large array of available phenotypes, using a hypothesis-free approach [18, 19]. The advantage of the hypothesis-free testing is the possible establishment of novel associations, reporting all the results, and not only the most statistically significant ones [20]. The threshold mainly used for statistical significance is the Bonferroni correction. It is stringent, since the phenotypes under investigation are considered to be independent, which is usually not the case [21]. Despite this, the large sample size used for the analysis counterweights for the multiple testing, leading to the replication of known associations and even the discovery of new ones [20].

We used PHEnome Scan Analysis (PHESANT) [19], which is a software package for performing comprehensive phenome scans in UK Biobank [22]. PHESANT tests the association of the exposure (independent variable), in our case the PRS of COVID-19, with quantitative and disease phenotypes in UK Biobank (dependent variables), using regression models. In more detail, PHESANT is implemented in statistical and computing software package, R [23]. A novel rule-based algorithm is used to categorize the phenotypes and the types of regressions, adjusted for the confounders. Based on these, the output of the software is result files depending on each test type: linear, logistic, ordered logistic and multinomial regressions. In addition, the results include a log-file providing information about the processing flow for each field and a model-fit-log file that shows model fit output for categorical unordered models. Lastly, the variable-flow-counts-all file, which includes a set of counts denoting the number of variables reaching each point in the processing flow, for example whether the variable has been analysed or has been excluded, or if some categories from the categorical fields have been excluded, are also provided in the results. In addition, for the continuous data, the inverse normal rank transformation is used to make certain they are normally distributed. To account for multiple testing, Bonferroni corrected threshold is calculated.

In order to assess the associations of the PRS with Hospital Episode data and data from the General practice (GP), we used the PheWAS library [18] in R [23]. This library is used to test the association of the exposure, the PRS of COVID-19 (independent variable), against each one of the phecodes (dependent variables), The software uses a function to convert the ICD-10 codes to ‘PheWAS codes’ or phecodes, which represent around 1,600 hierarchical phenotypes formed from grouped ICD-10 codes. In total, the phecodes are divided into 17 distinct categories, such as circulatory system, endocrine/metabolic, mental disorders, neurological, respiratory, plus 1 unlabelled category (denoted as NULL). Next, binary logistic regression models are employed to examine the association of the trait of interest with each phecode, adjusted for the confounders. For our analysis, Bonferroni corrected threshold is applied.

## Study Population

We examined the UK Biobank, a prospective cohort of 502,504 participants, aged 37-73 years old, recruited between 2006 and 2010. The dataset we used includes a variety of phenotypes including blood measurement, clinical assessments, anthropometry, cognitive function, hearing, arterial stiffness, hand grip strength, spirometry, ECG, data on cancer and death registries, health and lifestyle medical conditions, operations, mental health, sociodemographic factors, lifestyle, family history, psychosocial factors and dietary intake [20]. The analysis was performed using the maximum of 379,655 White British participants, for the variables that had no missing values.

In the present analysis, as trait of interest (exposure) we used the PRS of COVID-19. The PRS is created based on the unweighted sum of increasing risk COVID-19 alleles an individual carries, based on data from the HGI COVID-19 GWAS [24].

For our analysis we used summary statistics from the A2, B2 and C2 analyses. For A2 analysis the phenotype is defined as “Very severe respiratory confirmed COVID-19 vs. population”. For analysis B2, the phenotype is defined as “Hospitalized COVID-19 vs. population”. For the C2 analysis the phenotype is defined as “COVID-19 vs. population”. Our analyses were focused on individuals of European ancestry (White British).

A higher allele score corresponds to more severe COVID-19, depending on the definition used for the GWAS meta-analysis defined above, and it is standardized to have mean of 0 and standard deviation of 1.

## SNP selection for the PRS

We used conditional and joint analysis (GCTA-COJO) [25, 26], is employed to select the COVID-19-associated Single Nucleotide Polymorphisms - SNPs for the PRS. A stepwise procedure is used for SNP selection and the joint effects of all selected SNPs are estimated after the model has been optimized. These SNPs are still genome-wide significant, independent and the variance explained by them is larger than considering only the leading SNP at each locus. In more detail, conditional analysis was performed using 50,000 unrelated and randomly sampled European participants from UK Biobank as LD reference. We performed analyses using P-value at p=5*10^−8^ to declare a genome-wide significant hit. Also, SNPs having allele frequency differences larger than 0.2, were excluded from the analysis along with SNPs having MAF ≤ 0.001. As adjustments we used age at recruitment, sex, genotype batch, to account for the model variability. We also adjusted for the first 15 genetic principal components PCs to control for confounding via population stratification.

The number of SNPs reported from each analysis are presented in Table 1.

**Table 1:** Analysis and the corresponding number of SNPs used in the PRS.

| Analysis | Number of SNPs from COJO analysis – $p < 5*10^{-8}$ |
| --- | --- |
| Very severe respiratory confirmed COVID-19 vs. population –A2 | 11 |
| Hospitalized COVID-19 vs. population –B2 | 9 |
| COVID-19 vs. population –C2 | 5 |

## Phenotypes

We used phenotypic data from the UK Biobank including data from Hospital Episode Statistics (HES) and General Practice (GP). HES is a database containing details of all admissions at NHS hospitals in England. All clinical data are coded using International Classification of Disease (ICD) codes and all operations and procedures are coded using the Office of Population Censuses and Surveys (OPCS); Classification of Surgical Operations and Procedures (4th revision). The most recent ones are ICD-10, which are the ones we used, including 11,726 unique ICD-10 codes for 410,321 unique participants.

GP data contain the vast majority of a person’s health care information; 40% of GP records are linked to UK Biobank cohort up to now. Multiple clinical coding data are provided using Read codes; a standard vocabulary for clinicians to record patient findings and procedures. The most recent is version 3 CTV3 or Read v3), which is the one we used. Regarding the participants, 999 were excluded because the clinical event or prescription date precedes participant date of birth and 66 were removed because the event date is a date in the future and is presumed to be a place-holder or other system default) [27]. However, the 80,982 unique Read v3 codes, were converted to ICD-10 codes [28], in order to be subsequently used from the PheWAS library [29] in R [23]. As a result, 9,492 unique ICD-10 codes for 161,333 unique participants.

## Results

PheWAS analyses were conducted to evaluate the effect of genetically determined risk for three COVID-19 outcomes, severe, hospitalised and general COVID-19 infection (see Methods), on a comprehensive set of phenotypes including disease in UK Biobank.

Severe respiratory confirmed COVID-19 vs. population (A2) PRS was significantly associated with increased levels of lymphocytes for both counts and percentage (beta = 0.013, P = 3.15e^-15^; beta = 0.01, p = 5.6 e^-10^), respectively. Furthermore, the PRS was associated with decreased levels of platelet count (beta = -0.008, p = 2.6e^-07^). Severe COVID-19 outcome captured by the genetic score was associated with lower sex hormone binding globulin (SHBG) (beta = -0.0075, p = 2.59e^-06^), as well as with decreased FEV1/FVC ratio (beta = -0.009, p = 1.8 e^-06^). In addition, severe COVID-19 was associated with lower levels of insulin like growth factor-1 (IGF-1) (beta = -0.010, P = 3.2e^-10^) as shown in Figure 1. For the HES data, none of the phenotypes passed the Bonferroni correction threshold of statistical significance (p=3.35e-^05^); most significant associations were with hemopoietic traits especially congenital coagulation defects (OR = 0.85, p=2.68 e^-04^). Similarly, for the GP data, none of the phenotypes passed the Bonferroni test.

**Figure 1:**
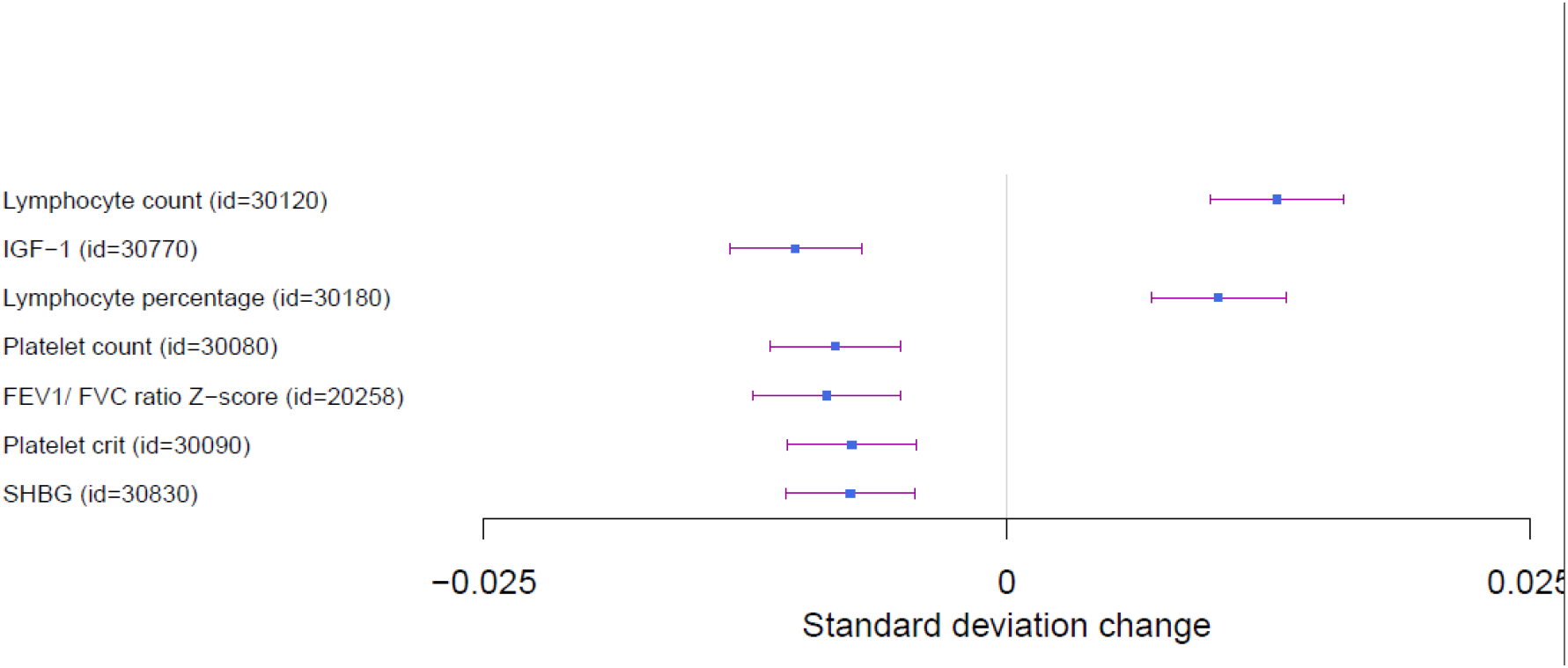
Forest plot of PheWAS analyses of Severe COVID-19 risk

Hospitalized COVID-19 vs. population PRS was associated with decreased levels of platelets counts (beta = -0.01, P = 1.2 e^-08^), and with increased lymphocyte counts (beta =0.007, p = 8.19e^-06^). Furthermore, we observed an association with decreased FEV1/FVC ratio (beta = -0.01, P = 1.4e^-08^) and increased body fat percentage (beta = 0.0057, p=2.6 e^-06^), especially with increased arms and trunk fat mass and percentages (beta = 0.003, p = 1.56 e^-06^), and decreased standing height (beta = - 0.006, p = 3.85 e^-07^), and sitting height (beta = -0.006, p = 3.38e^-07^) (Figure 2). No association signal passed Bonferroni correction in neither the HES nor GP data analysis.

**Figure 2:**
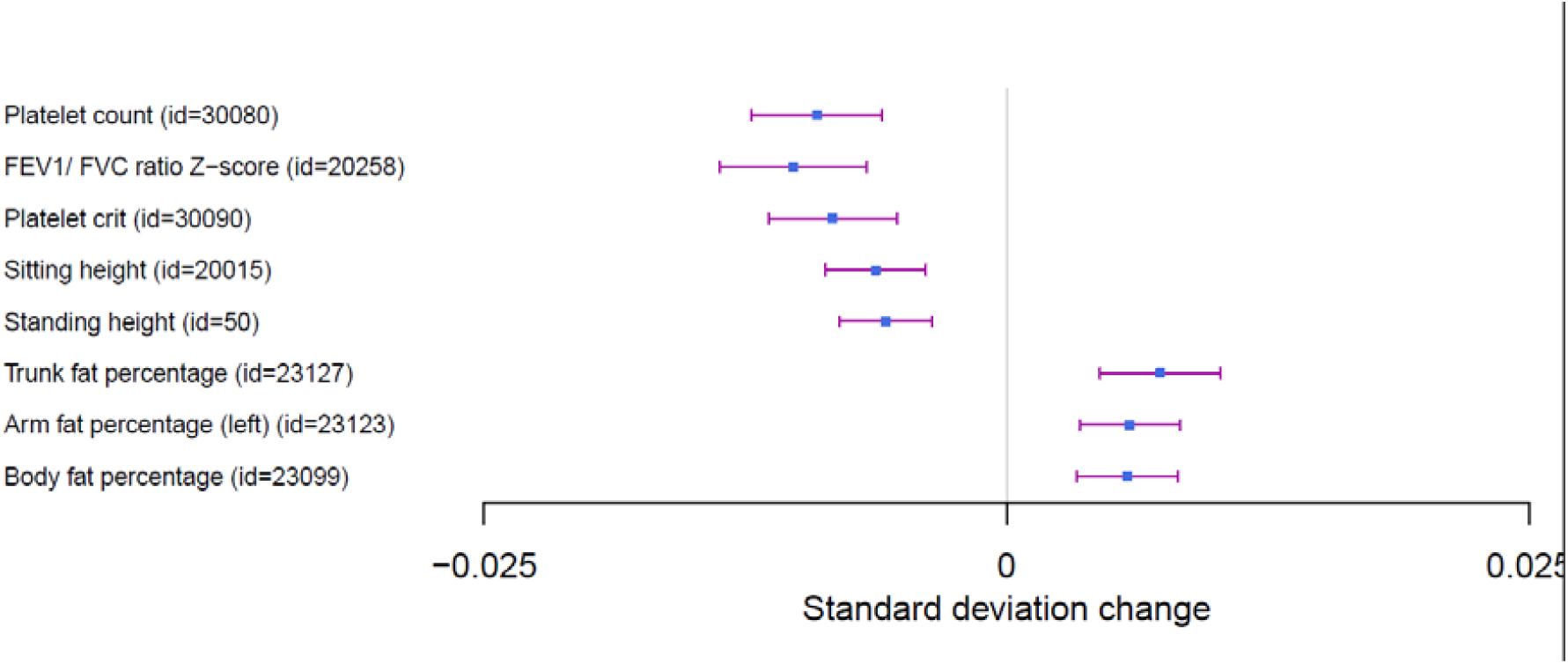
Forest plot of PheWAS analyses of Hospitalised COVID-19 risk

The general COVID-19 infection vs population (C2) PRS was associated with lower levels of alkaline phosphate (beta = -0.084, p<2.23 e^-308^) and aspartate aminotransferase (beta = -0.01, p=4.32e^-09^). Contrary to aspartate aminotransferase, a positive association was found with alanine aminotransferase (beta = 0.01, p = 9.41 e^-12^).

Finally, as shown in Figure 3, several haematological traits showed a significant association including lower levels of monocyte counts (beta = -0.02, p=1.81e^-31^), RBC cells (beta = -0.01 p=1.25e^-14^), neutrophils (beta = -0.01, p=1.8e^-10^) and lymphocytes (beta=0.01, p=6.68e^-09^). Moreover, we identified significant signals between COVID-19 susceptibility with blood clots in the leg (OR= 1.1, p = 1.66e^-16^) and with increased risk for blood clots in the lung (OR = 1.12, p = 1.45 e^-10^).

**Figure 3:**
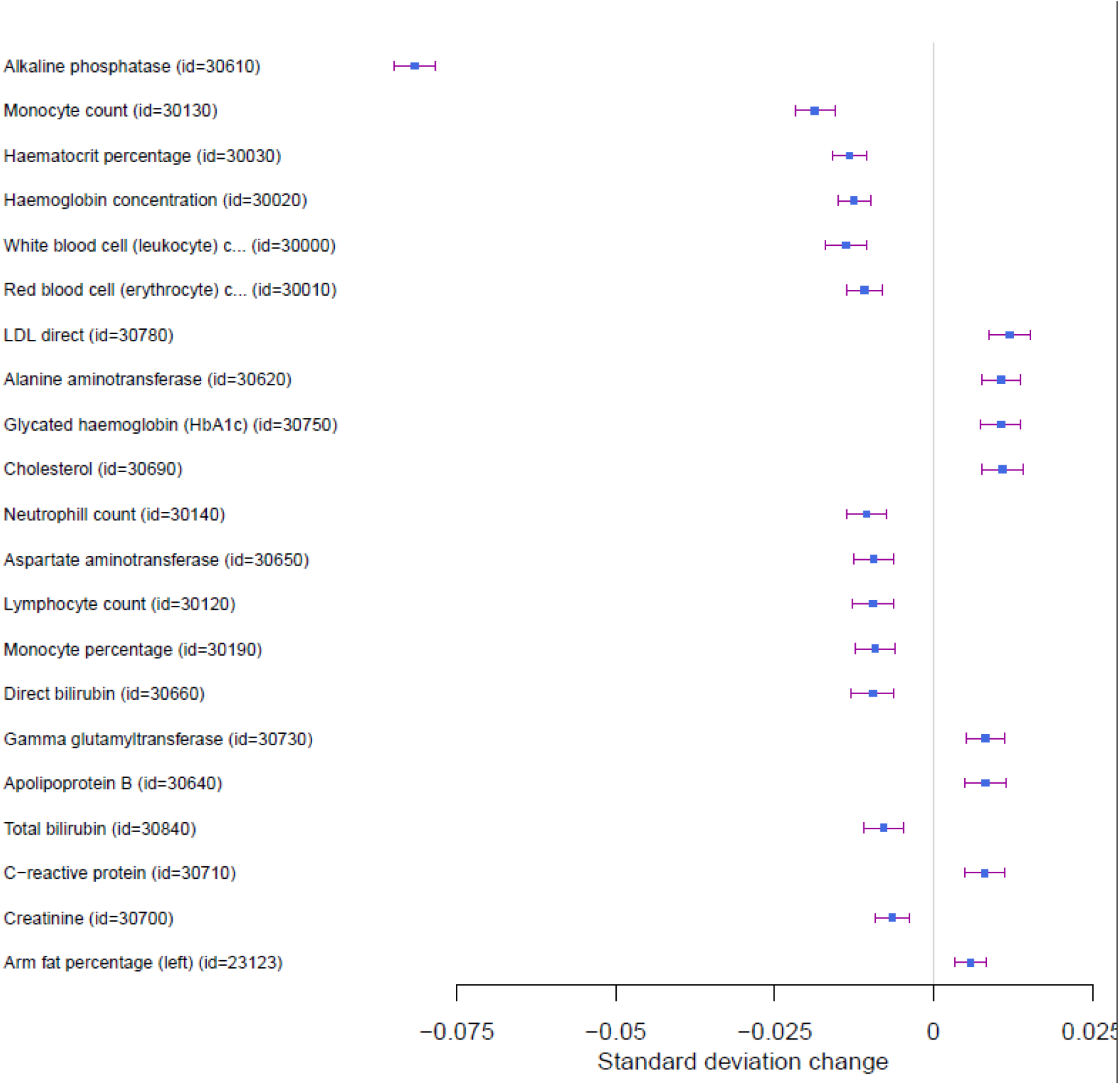
Forest plot of Phewas analyses of general COVID-19 susceptibility.

For the HES data, COVID-19 risk was associated with disorders of circulatory system were (OR = 1.04, p = 1.31e^-09^) (Figure 4A). Furthermore, in the GP data, COVID-19 risk was shown to be associated with increased risk for phlebitis and thrombophlebitis (OR = 1.11, p = 5.36e^-08^), phlebitis and thrombophlebitis of lower extremities (OR = 1.15, p = 3.43e^-05^) (Figure 4B). Full results for all the analyses are provided in Supplementary Data Files.

**Figure 4:**
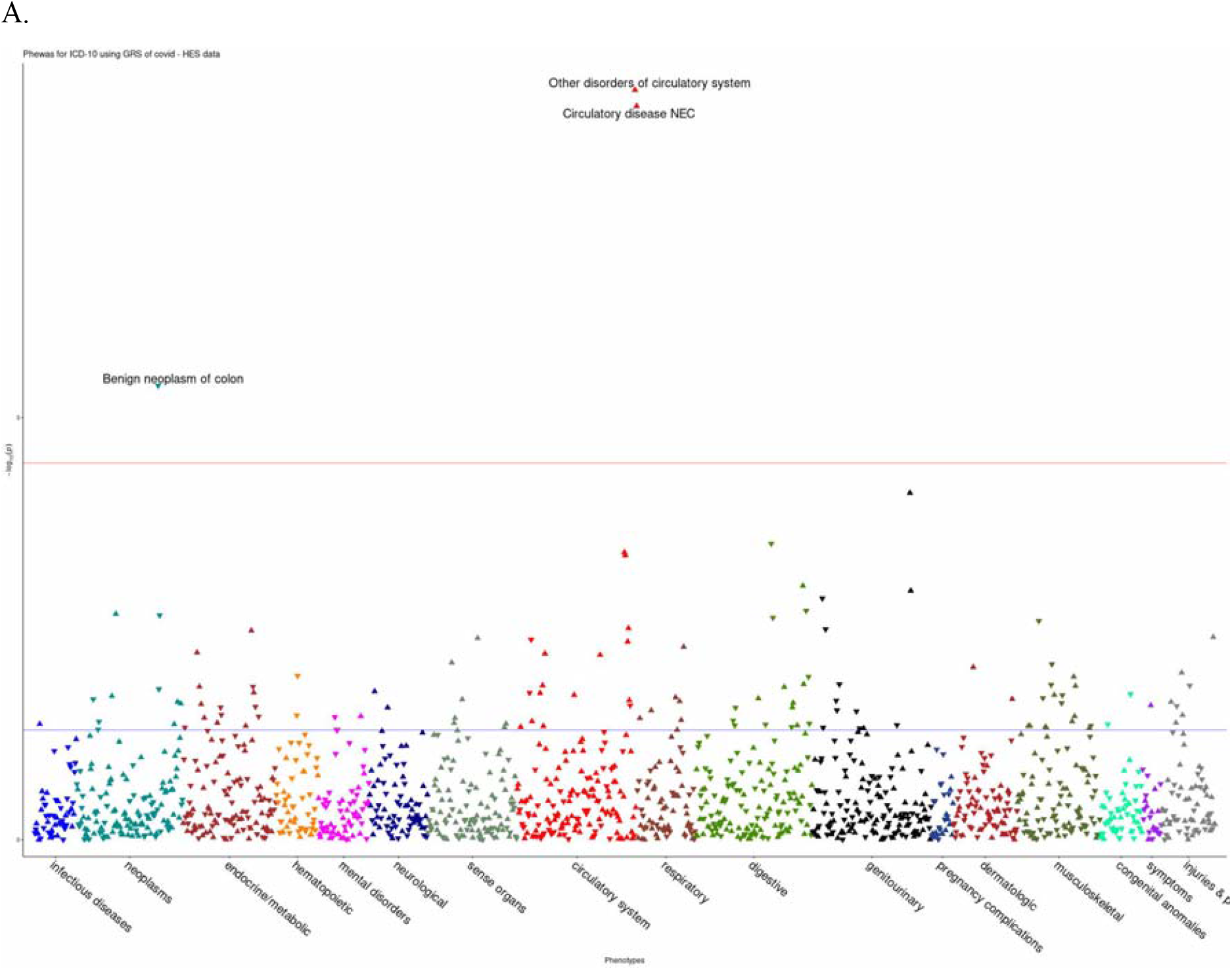
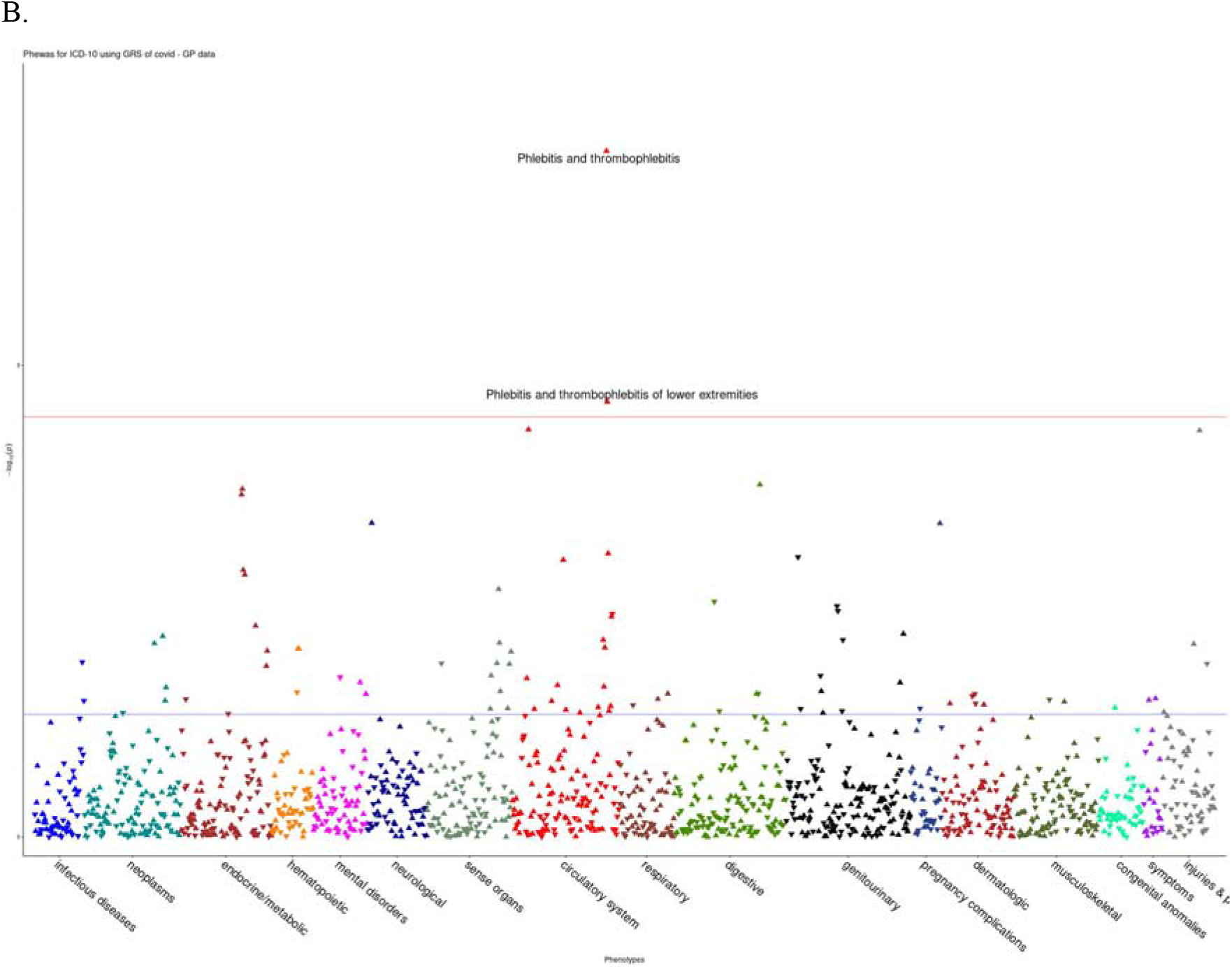
PheWAS associations for COVID-19 susceptibility in HES (A) and GP (B) data.

## Discussion

Our PheWAS analyses in UK Biobank identified a spectrum of associations between both genetically determined COVID-19 susceptibility and severity and a number of traits. Severe COVID-19 outcome was associated with increased counts and percentages of lymphocytes. However, several studies have also reported an association of lymphopenia with COVID-19 severity, in accordance with our findings for general COVID-19 susceptibility [30]. Lymphopenia is rarely observed in children infected with COVID-19, a group in which mortality rate is very low. Lymphocytes are known to play a crucial role in viral infections. Furthermore, atypical reactive lymphocytes are present in COVID-19 patients with better prognosis [31]. However, additional studies are needed to fully delineate the pathophysiology of immunological cells contribution to the disease.

Severe COVID-19 risk was associated with lower platelet count, in agreement with meta-analytic studies [32]. Thrombocytopenia has been shown to be common in severe cases of COVID-19 during infection, proposing it as a clinical indicator for illness progression during hospitalisation [33]. Those who are severely affected have shown coagulation abnormalities and clotting complications such as microvascular events, which have been commonly reported in retrieved tissues from severe cases [34]. Furthermore, the presence of platelet aggregates and macro-thrombocytes have been associated with COVID-19 patients admitted to ICU [35]. In a study conducted by Maine et al [36], they reported an increased expression of platelet genes resulting in an increased activation and aggregation in patients infected by coronavirus reflecting their hyperactive status [36].

SARS-CoV-2 has been associated with abnormal pulmonary function, mainly by causing severe acute respiratory syndrome and being a significant reason for mortality and hospitalisation [37]. In this study, increased risk of severe COVID-19 and hospitalisation were associated with decreased FEV/FVC z-score. Additionally, studies have noted post-recovery changes resembling restrictive pulmonary diseases and variations in resorting to normal range levels [38, 39].

We also observed that genetically determined severe COVID-19 was associated with lower levels of IGF-1. Fan et al (2021) suggested that a higher concentration of IGF-1 has an association with a lower risk of the mortality of COVID-19 [40]. Although IGF-1 role in severe respiratory distress has not been fully delineated, further research and understanding of its pathway and contribution shows potential as a therapeutic mean. Severe COVID-19 risk was associated with decreased levels of SHBG. SHBG has a major role in balancing testosterone levels as it has a greater affinity for it more than oestrogen[41]. Interestingly, most male ICU admitted patients with COVID-19 had shown lower testosterone levels, deeming it a screening parameter for early detection of high-risk patients [42, 43]. Nevertheless, current studies lack describing the role of sex hormones and their binding proteins in COVID-19.

The risk of COVID-19 hospitalisation risk outcome was associated with increased trunk and arms fat mass and lower standing and sitting height. Several studies have shown that most hospitalised COVID-19 patients are obese [44]. Furthermore, severe outcome of COVID-19 has been associated with high visceral fat adiposity in European adults [45]. Obesity is identified as abnormal secretions of cytokines and adipokines resulting in increased inflammation; thus, it might contribute to the outcomes requiring hospitalization [46, 47]. Although abnormal liver enzymes were noted in the majority of COVID-19 patients, studies were not conclusive on the significance of liver function enzymes being informative predictors of the severity of the disease progression [48, 49].

For general COVID-19 susceptibility, we observed significant associations with increased phlebitis and thrombophlebitis. This result is in agreement with the available meta-analytic literature as COVID-19 procoagulant activity such as deep vein thrombosis, pulmonary embolism, and superficial phlebitis, are prevalent in moderate to severe cases [50]. Furthermore, we identify significant association of general COVID-19 with increased blood clot events in leg and lungs. In a meta-analytic study blood clots rates of COVID-19 have shown to be a risk of mortality [51].

The reported associations between both genetically determined COVID-19 susceptibility and severity and other diseases adds to the identification and stratification of individuals at increased risk. Further work will be required to better understand the biological and clinical value of these findings including long-term complications after infection.

## Data Availability

publically available

## Acknowledgments

The Barts Biomedical Research Centre funded by the UK National Institute for Health Research (NIHR) has supported COVID related research.

UK Biobank participants (Application 53723)

